# Prediction of individuals at high-risk of chronic kidney disease during treatment with lithium for bipolar disorder

**DOI:** 10.1101/2020.12.31.20248928

**Authors:** Joseph F Hayes, David PJ Osborn, Emma Francis, Gareth Ambler, Laurie A Tomlinson, Magnus Boman, Ian Wong, John Geddes, Christina Dalman, Glyn Lewis

**Affiliations:** Division of Psychiatry, UCL; Camden and Islington NHS Foundation Trust; Department of Statistical Science, UCL; Department of non-Communicable Disease Epidemiology, LSHTM; Division of Software and Computer Systems, School of Electrical Engineering and Computer Science, KTH; Department of Learning, Informatics, Management and Ethics, Karolinska Institute; Centre for Safe Medication Practice and Research, Department of Pharmacology and Pharmacy, The University of Hong Kong; Research Department of Practice and Policy, School of Pharmacy, UCL; Department of Psychiatry, University of Oxford; Department of Global Public Health, Karolinska Institute

## Abstract

**Background:** Lithium is the most effective treatment in bipolar disorder. Its use is limited by concerns about risk of chronic kidney disease (CKD). We aimed to develop a model to predict risk of CKD following lithium treatment initiation, by identifying individuals with a high-risk trajectory of renal function.

**Methods:** We used United Kingdom Clinical Practice Research Datalink (CPRD) electronic heath records (EHRs) from 2000-2018. CPRD Aurum for prediction model development and CPRD Gold for external validation. We used elastic net to generate a prediction model from potential features. We performed discrimination and calibration assessments in an external validation data set.

We included all patients aged ≥16 with bipolar disorder prescribed lithium. To be included patients had to have ≥1 year of follow-up before lithium initiation, ≥3 estimated glomerular filtration rate (eGFR) measures after lithium initiation (to be able to determine a trajectory) and a normal (≥60 mL/min/1.73m^2^) eGFR at lithium initiation (baseline). In the Aurum development cohort 1609 fulfilled these criteria. The Gold external validation cohort included 934 patients.

We included 44 potential baseline features in the prediction model, including sociodemographic, mental and physical heath and drug treatment characteristics. We compared a full model with the 3-variable five-year kidney failure risk equation (KFRE) and a 3-variable elastic net model.

We used group-based trajectory modelling to identify latent trajectory groups for eGFR. We were interested in the group with deteriorating renal function (the high-risk group).

**Findings:** The high-risk group included 191 (11.87%) of the Aurum cohort and 137 (14.67%) of the Gold cohort, of these 168 (87.96%) and 117 (85.40%) respectively developed CKD 3a or more severe during follow-up. The model, developed in Aurum, had a ROC area of 0.879 (95%CI 0.853-0.904) in the Gold external validation data set. At the empirical optimal cut-point defined in the development dataset, the model had a sensitivity of 0. 91 (95%CI 0.84-0.97) and a specificity of 0.74 (95% CI 0.67-0.82). However, a 3-variable elastic net model (including only age, sex and baseline eGFR) performed similarly well (ROC area 0.888; 95%CI 0.864-0.912), as did the KFRE (ROC area 0.870; 95%CI 0.841-0.898).

**Conclusions:** Individuals at high-risk of a poor trajectory of renal function can be identified before initiation of lithium treatment by a simple equation including age, sex and baseline eGFR. We did not identify strong predicters of renal impairment specific to lithium treated patients.

## Background

Lithium is the most effective maintenance treatment for bipolar disorder, and is first-line in all international clinical practice guidelines (1). However, its use has been declining globally (2). Reasons for this include the required monitoring due to its narrow therapeutic window and concerns about adverse effects, particularly irreversible renal failure (3). In fact, renal failure is rare (4), with end-stage renal failure occurring at similar rates to those treated with other mood stabilisers (5), but more commonly than the general population (0.23% vs 0.11% (6)). Bipolar disorder itself appears to be associated with increased risk of renal failure independent of lithium exposure (7). There are inconsistencies in the existing literature about the association between renal failure and lithium treatment duration and episodes of lithium toxicity (8).

Being able to identify individuals at risk of compromised renal function would have high clinical utility; it would encourage the use of this effective treatment in those at low-risk and so improve outcomes for people with bipolar disorder. In the general population, established risk factors for CKD include age, sex, ethnicity, family history, smoking, obesity, hypertension, diabetes mellitus, excessive alcohol consumption and acute kidney injury (9). Prediction models have been developed for end stage renal failure in groups with a range of underlying risk (10-14). These tend to include a small number of core features including age, gender, ethnicity, eGFR and albuminuria. Models then vary in terms of additional features such as glucose, blood pressure, haemoglobin, lipids, calcium and phosphate. It is unclear if features related to mental health are useful in predicting CKD risk at the point of lithium initiation. It is also likely that risk factors for CKD cluster differently in patients with bipolar disorder prescribed lithium [8] so we cannot assume that risk prediction models for CKD that are of value in the general population would apply to people with bipolar disorder receiving lithium. Because CKD requiring clinical intervention (CKD stage 4 or more severe – eGFR<30 mL/min/1.73m^2^) is a rare and late-stage outcome we aimed to develop a model to classify individuals into high-risk and low-risk trajectories of renal function following lithium treatment initiation.

## Methods

### Population

This study used patient data from the Clinical Practice Research Datalink (CPRD) Gold and Aurum databases between 1 January 2000 and 31 December 2018. CPRD contains electronic health records (EHRs) from general practices across the United Kingdom. Combined, these databases include 42 million patient records from over 1,800 primary care practices (www.cprd.com). Both databases contain coded and anonymised data including demographic details, symptoms, diagnoses, prescribed medication, laboratory tests and referrals. CPRD Gold contains data contributed by practices using Vision software and CPRD Aurum contains data contributed by practices using EMIS Web software (15, 16). Contributing practices have different geographical distributions; CPRD Gold contains patients from the whole of the UK, whereas Aurum contains only practices from England and Northern Ireland. Therefore, there are some differences in population structures. We used data from the Aurum database for development of our prediction model and data from the Gold database for external validation. Ethical approval for this study was obtained from the Independent Scientific Advisory Committee of CPRD (protocol no. 18_316). Informed consent was waived because data are anonymised for research purposes. In line with ethical guidance subgroups containing fewer than 5 people are censored in the results section.

### Cohort definition

The cohort comprised any patient who; was aged 16 or over, ever received a diagnosis of bipolar disorder in their clinical record, was prescribed lithium (defined as receiving two or more concurrent prescriptions), had at least a year of follow-up before their first lithium prescription and no previous record of being prescribed lithium (to capture patients’ first exposure to lithium), had at least three estimated glomerular filtration rate (eGFR) measures after lithium initiation and had a baseline measure of eGFR ≥60 mL/min/1.73m^2^ before starting lithium (normal or close to normal renal function).

### Renal function trajectories

eGFR values were calculated from recorded creatinine blood tests using the CKD-EPI equation (17). Using the eGFR, and the date the blood test was performed relative to lithium initiation, we conducted group-based trajectory modelling to identify latent subgroups within the cohort (18). We included lithium exposure as a time-varying covariate, as rate of change in eGFR may potentially differ between the lithium exposed period and following lithium cessation. In the process of determining the number of trajectory groups, we initially used a cubic polynomial function for all groups. The final number of groups was determined based on the Bayesian Information Criterion (BIC), trajectory shapes for similarity, and the proportion of cohort members in each class (19). We initially set a 2-group model and increased the number of groups until BIC was minimised, but no group was less than 10% of the total cohort. After identifying the optimal number of groups, the level of the polynomial function for each group was reduced from cubic to zero-order until the BIC was minimised. With this final model, each participant was assigned to one of the subgroups based on maximum posterior probability. We were primarily interested in the group predicted to have the most rapidly declining eGFR trajectory; referred to as the high-risk group.

### Prediction model features

We identified features present in a patient’s record before they commenced lithium treatment as potential predictors of being in the high-risk group. These included predictors of impaired renal function in the general population and features related to mental health and its treatment:

#### Sociodemographics

Age, sex, ethnicity (as Black, Asian and Minority Ethnic (BAME) vs White), relationship status (single vs. in a relationship).

#### Mental health characteristics

Illness duration before lithium initiation, presentations for depression, mania, anxiety, psychosis, stress, self-harm, disturbed sleep.

#### Physical health characteristics

Hyper/hypocalcaemia, hypo/hyperthyroidism, high LDL cholesterol, low HDL cholesterol, hypertension, coronary heart disease, a measure of eGFR<60 mL/min/1.73m^2^ any time before lithium initiation, Type II Diabetes Mellitus, asthma, weight loss, peptic ulcer, iron deficiency anaemia, liver disease, chronic pulmonary disease, neurological disorders.

#### Health behaviours

Smoking status (never smoked, current smoker, ex-smoker), body mass index group (underweight, healthy weight, overweight, obese), cannabis use, other substance misuse, alcohol misuse.

#### Other drug treatment

Antipsychotic prescription, other mood stabiliser prescription, antidepressant prescription.

#### Interactions

Baseline eGFR with sex and age, sex with age and body mass index group.

### Statistical Analysis

We described differences in prevalence of binary covariates and medians of continuous covariates between high-risk and low-risk groups using p-values from chi-squared tests. We used probit elastic net regression with 10-fold cross-validation to perform variable selection and penalization of coefficients to generate the prediction model in the Aurum cohort. Elastic net is a regularization method for regression and classification models which comprises the Least Absolute Shrinkage And Selection Operator (LASSO) penalty (L1) and the ridge penalty (L2)(20). The LASSO (L1) penalty function performs variable selection and dimension reduction by shrinking coefficients, while the ridge (L2) penalty function shrinks the coefficients of correlated variables toward their average. The overall elastic net is a function of parameters λ and α (0 ≤ α ≤ 1), with λ being a parameter for the level of penalty, while α being the weight of L1 penalty and (1-α) being that of L2 penalty function. We reported Receiver Operating Characteristic (ROC) area (and 95% confidence interval)(CI), sensitivity and specificity at the empirical optimal cut-off point using Youden’s index and the predictive accuracy. We compared the derived full model with predictions from the 3-variable five year kidney failure risk equation (KFRE) which includes age, sex and eGFR, and an elastic net model containing only these 3 variables (14). We chose the 3-variable KFRE as albumin-to-creatine ratio was poorly recorded before lithium initiation and the 3-variable model performed well in previous validation studies (ROC area 0.79)(21).

### External validation

We used patient data from CPRD Gold for external validation of the model generated in the Aurum cohort. To categorise individuals at high-risk of a rapid decline in eGFR, we ran group-based trajectory models of the eGFRs independently of the Aurum patients’ trajectory model. We compared trajectory group membership with the predicted group membership from the Aurum model. We reported the ROC area, sensitivity and specificity at the cut-off point defined in the development data, brier score, predictive accuracy, calibration belt (a graphical approach designed to evaluate the goodness of fit of binary outcome models) (22) and decision curve analysis. We examined how well the model could predict CKD stage 3b or more severe (eGFR<45 mL/min/1.73m^2^) during follow-up. We also compared the full model with simple models: the 3-variable KFRE and 3-variable elastic net.

### Post hoc supplementary analysis

We combined data from the Aurum and Gold datasets for patients who initiated lithium treatment with a baseline eGFR ≥90 mL/min/1.73m^2^. We adopted the same approach in this smaller cohort: we identified a high-risk trajectory group using group-based trajectory modelling and then used the full model, 3-variable KFRE and 3-variable elastic net to predict group membership. We also examined how well this model could predict CKD stage 3a or more severe (eGFR<60 mL/min/1.73m^2^) during follow-up. This analysis was completed to address issues arising from the strong association between baseline eGFR and future eGFR measurements in the initial model. All analysis was completed using Stata 16 (23).

## Findings

We identified 1609 patients in the development sample (Aurum cohort), with a median of 14 (IQR 7-26) eGFR test results each. The median length of lithium treatment was 1.42 years (IQR 0.53-3.58). Of these patients, 401 (24.92%) developed CKD stage 3a or more severe (eGFR<60 mL/min/1.73m^2^), 38 (2.36%) CKD stage 3b (eGFR<45 mL/min/1.73m^2^), but none developed CKD stage 4. In total, 158 (9.82%) died during follow-up.

To categorize risk groups based on eGFR trajectories we chose a 5-group model, all groups with cubic trajectories (BIC=3566.99). This defined 11.87% of the cohort as high-risk. Models with 6 groups had lower BIC but included one group with less than 10% of the cohort.

Trajectories of the high-risk vs other groups (combined group 2-5) are shown in Figure 1 and described in Table 1. Of those in the high-risk group 168 (87.96%) develop CKD stage 3a or more severe, and 25 (13.09%) developed stage 3b or more severe, compared to 233 (16.43%) and 13 (0.92%) respectively in the low-risk group.

**Table 1.** Characteristics of lithium prescribed patients by risk group in Aurum

| Feature | High-risk | Low-risk | P-value |
| --- | --- | --- | --- |
| **Patient characteristics** |  |  |  |
| Total, N (%) | 191 (11.87) | 1418 (88.13) |  |
| Female, n (%) | 178 (93.19) | 852 (60.08) | <0.001 |
| Age, median (IQR) | 41.58 (32.14-50.72) | 45.80 (36.39-56.59) | <0.001 |
| BAME, n (%) | 7 (3.66) | 78 (5.50) | 0.287 |
| In a relationship, n (%) | 32 (16.75) | 240 (16.93) | 0.953 |
| Death during follow-up | 5 (2.62) | 153 (10.79) | <0.001 |
| **Lithium exposure characteristics** |  |  |  |
| Lithium treatment duration (years), median (IQR) | 1.66 (0.72-4.11) | 1.51 (0.59-3.85) | 0.482 |
| Ever lithium toxic (>1.5mmol/L), n (%) | 5 (2.62) | 62 (4.37) | 0.256 |
| Follow-up after stopping lithium (years), median (IQR) | 2.92 (0-7.14) | 4.07 (0.24-9.43) | 0.064 |
| **Renal function characteristics** |  |  |  |
| eGFR tests during follow-up, median (IQR) | 12 (7-20) | 14 (8-27) | 0.004 |
| Baseline eGFR (mL/min/1.73m<sup>2</sup>), median (IQR) | 67 (63-72) | 81 (72-91) | <0.001 |
| Developed CKD stage 3a or more severe (eGFR<60 mL/min/1.73m<sup>2</sup>) | 168 (87.96) | 233 (16.43) | <0.001 |
| Developed CKD stage 3b or more severe (eGFR<45 mL/min/1.73m<sup>2</sup>) | 25 (13.09) | 13 (0.92) | <0.001 |
| **Pre-lithium mental health characteristics** |  |  |  |
| Depression, n (%) | 160 (83.77) | 1167 (82.30) | 0.616 |
| Anxiety, n (%) | 60 (31.41) | 403 (28.42) | 0.391 |
| Psychosis, n (%) | 33 (17.28) | 266 (18.76) | 0.621 |
| Stress, n (%) | 45 (23.56) | 262 (18.48) | 0.093 |
| Self-harm, n (%) | 39 (20.42) | 217 (15.30) | 0.070 |
| Disturbed sleep, n (%) | 48(25.13) | 315 (22.21) | 0.365 |
| Illness duration (years), median (IQR) | 8.47 (4.03-15.65) | 8.45 (2.86-16.30) | 0.931 |
| **Pre-lithium physical health characteristics** |  |  |  |
| Hypertension, n (%) | 33 (17.28) | 260 (18.34) | 0.722 |
| Migraine, n (%) | 40 (20.94) | 138 (9.73) | <0.001 |
| Type II diabetes mellitus, n (%) | 18 (9.42) | 107 (7.55) | 0.363 |
| Thyroid disease |  |  |  |
| Hypothyroidism, n (%) | 5 (2.62) | 70 (4.94) | 0.154 |
| Hyperthyroidism, n (%) | * | 8 (0.56) | 0.113 |
| Calcium abnormalities |  |  |  |
| hypocalcaemia, n (%) | * | 12 (0.85) | 0.640 |
| hypercalcaemia, n (%) | * | 13 (0.93) | 0.184 |
| Cholesterol abnormalities |  |  |  |
| High LDL, n (%) | 47 (24.61) | 264 (18.62) | 0.049 |
| Low HDL, n (%) | 16 (8.38) | 101 (7.12) | 0.531 |
| Asthma, n (%) | 52 (27.23) | 282 (19.89) | 0.019 |
| Chronic obstructive pulmonary disease, n (%) | 57 (29.84) | 341 (24.05) | 0.081 |
| Anaemia, n (%) | 11 (5.76) | 67 (4.72) | 0.532 |
| Peptic ulcer, n (%) | 2 (1.05) | 20 (1.41) | 0.685 |
| Coronary heart disease, n (%) | 5 (2.62) | 68 (4.80) | 0.175 |
| Liver disease, n (%) | * | 23 (1.62) | 0.546 |
| Neurological disorders, n (%) | 15 (7.85) | 89 (6.28) | 0.405 |
| Rheumatoid arthritis, n (%) | 6 (3.14) | 28 (1.97) | 0.293 |
| Weight loss, n (%) | 8 (4.19) | 31 (2.19) | 0.091 |
| Ever eGFR<60 mL/min/1.73m<sup>2</sup>, n (%) | * | 40 (2.82) | 0.019 |
| **Health behaviours** |  |  |  |
| Smoking status, n (%) |  |  | 0.51 |
| Never smoked | 56 (29.32) | 403 (28.42) |  |
| Current smoker | 93 (48.69) | 581 (41.47) |  |
| Ex-smoker | 42 (21.99) | 427 (30.11) |  |
| Body mass index, n (%) |  |  | <0.001 |
| Underweight | * | 37 (2.61) |  |
| Healthy weight | 57 (29.84) | 616 (43.44) |  |
| Overweight | 50 (26.18) | 409 (28.84) |  |
| Obese | 80 (41.88) | 356 (25.11) |  |
| Cannabis use, n (%) | * | 14 (0.99) | 0.938 |
| Other substance misuse, n (%) | 10 (5.24) | 99 (6.98) | 0.367 |
| Alcohol misuse, n (%) | 10 (5.24) | 82 (5.78) | 0.760 |
| **Other psychiatric drug treatment** |  |  |  |
| Antipsychotic previously, n (%) | 104 (54.45) | 766 (54.02) | 0.911 |
| Mood stabiliser previously, n (%) | 72 (37.70) | 432 (30.47) | 0.043 |
| SSRI previously, n (%) | 50 (26.18) | 419 (29.55) | 0.336 |
| TCA previously, n (%) | 55 (28.80) | 334 (23.55) | 0.112 |
| Other antidepressant previously, n (%) | 43 (22.51) | 236 (16.64) | 0.044 |

\*n<5 individuals

**Figure 1.**
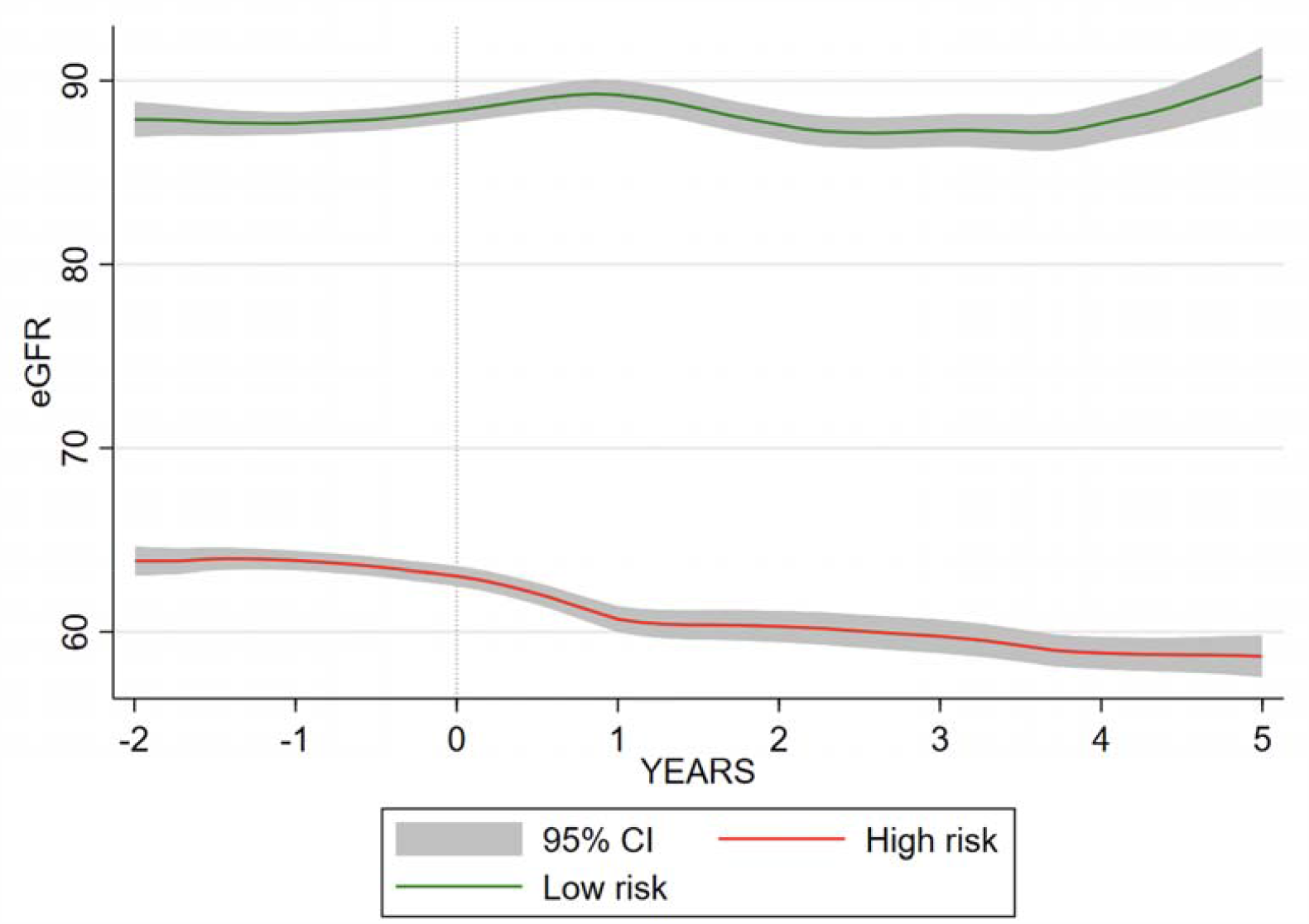
High-risk and low-risk eGFR trajectory in relation to end of lithium exposure in Aurum

Patients in the high-risk group were more likely to be female, younger at lithium initiation, have a lower eGFR before starting lithium and be obese. They were more likely to have a pre-existing diagnosis of migraine. They were more likely to have a record of high LDL cholesterol. They were less likely to have had an eGFR<60 mL/min/1.73m^2^ any time before baseline.

There was no statistical evidence of a difference in duration of lithium treatment and incidence of lithium toxicity (>1.5mmol/L) between groups. Those in the high-risk group were less likely to die during follow-up and had fewer eGFR tests in total.

We used 44 features known to the clinician prior to lithium initiation to generate a prediction model for being in the high-risk group. Elastic net with 10-fold cross-validation fitted a model with λ=0.014 and α=1.00. The ROC area 0.868 (95%CI 0.844-0.891) (Figure 2). The empirical optimal cut-point was 0.134 with a sensitivity of 0.86 (95%CI 0.78-0.94) and a specificity of 0.73 (95%CI 0.63-0.84). The Youden index was 0.589. This gave a prediction accuracy of 74.54% (Table 2).

**Table 2.** Prediction of high-risk group membership

| MODEL | ROC area (95%CI) | Sensitivity (95%CI) | Specificity (95%CI) | Accuracy % |
| --- | --- | --- | --- | --- |
| **Development (Aurum)** |  |  |  |  |
| Full | 0.868 (95%CI 0.844-0.891) | 0.86 (0.78-0.94) | 0.73 (0.66-0.80) | 74.54 |
| 3-variable KFRE | 0.828 (0.801-0.855) | 0.87 (0.77-0.96) | 0.64 (0.54-0.74) | 66.73 |
| 3-variable elastic net | 0.852 (0.827-0.856) | 0.86 (0.77-0.95) | 0.68 (0.58-0.78) | 70.14 |
| **Validation (Gold)** |  |  |  |  |
| Full | 0.879 (0.853-0.904) | 0.91 (0.84-0.97) | 0.74 (0.67-0.81) | 76.55 |
| 3-variable KFRE | 0.870 (0.841-0.898) | 0.86 (0.78-0.94) | 0.75 (0.68-0.83) | 76.66 |
| 3-variable elastic net | 0.888 (0.864-0.912) | 0.86 (0.79-0.94) | 0.80 (0.73-0.87) | 80.90 |

**Figure 2.**
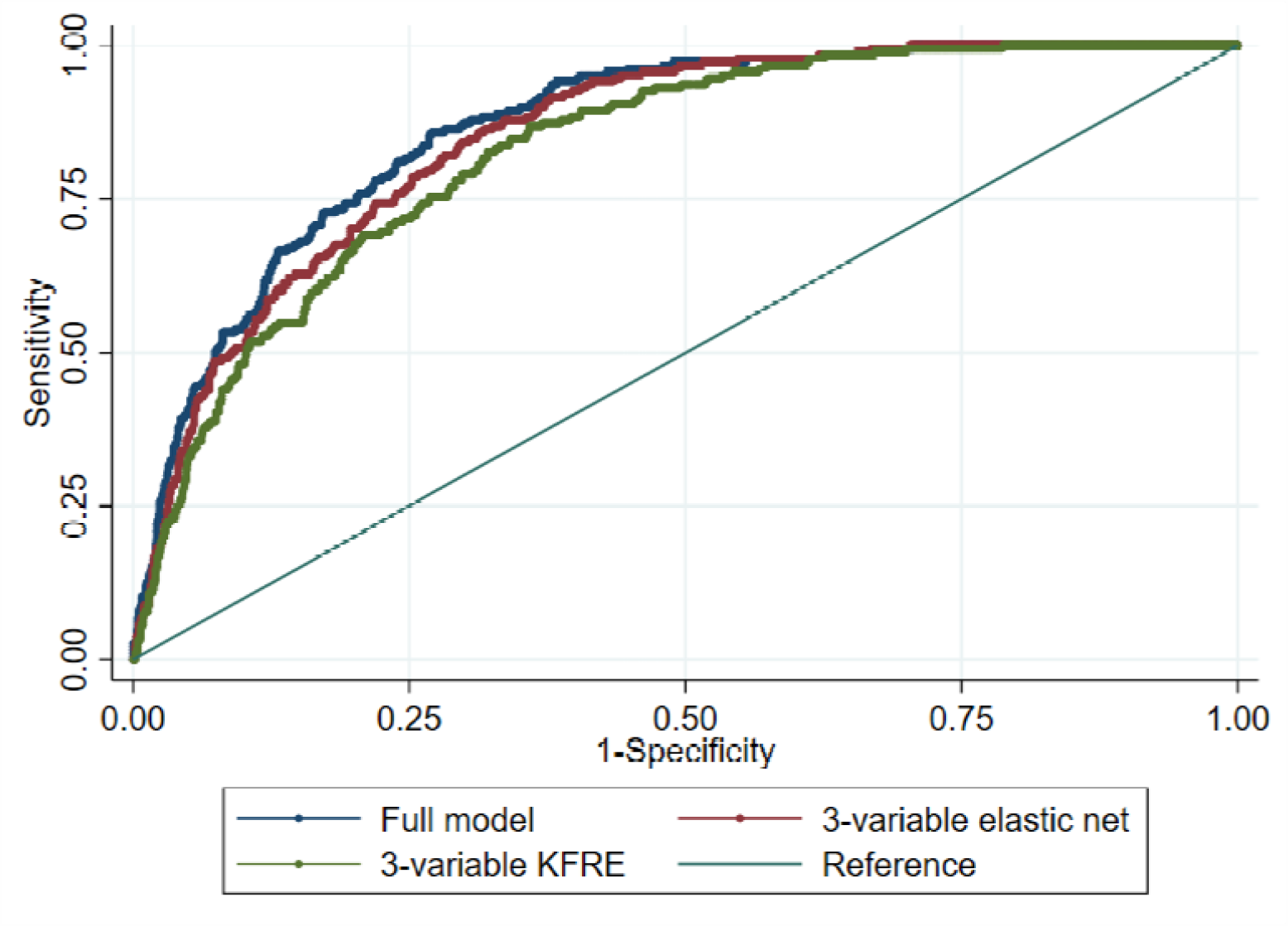
Sensitivity vs specificity of the high-risk trajectory prediction model in Aurum

Features retained in the model were (in order of coefficient size): baseline eGFR, sex, sex by BMI group interaction, baseline eGFR by age interaction, hypothyroidism, migraine, BMI group, SSRI exposure, high LDL cholesterol, BAME, hyperthyroidism, smoking status, type 2 diabetes mellitus, self-harm. The 3-variable KFRE and the 3-variable elastic net model performed similarly well to the full model: ROC area 0.828 (95%CI 0.801-0.855) and ROC area 0.852 (95%CI 0.827-0.876) respectively (Table 2).

### External validation

The external validation data set (Gold cohort) included 934 individuals. We developed new trajectory groups independently for these patients. BIC in the group-based trajectory model was minimised by a 5-group solution, with cubic or quadratic polynomials fitted for each group trajectory; 3, 2, 2, 3, 3 respectively from “highest risk” to “lowest risk” groups (BIC=1919.07). Of the total Gold cohort, 14.67% (n=137) were in the high-risk group. Of the total cohort, 229 (24.52%) developed CKD stage 3a or more severe and 14 (1.50%) CKD stage 3b or more severe.

Patient characteristics by risk group are described in Table 3, and trajectories relative to end of lithium exposure are shown in Figure 3. Of those in the high-risk group, 117 (85.40%) develop CKD stage 3a or more severe, and 14 (10.22%) developed stage 3b or more severe, compared to 112 (14.05%) and <5 respectively in the low-risk group.

**Table 3.** Characteristics of lithium prescribed patients by risk group in Gold

| Feature | High-risk | Low-risk | P-value |
| --- | --- | --- | --- |
| **Patient characteristics** |  |  |  |
| Total, N (%) | 137 (14.67) | 797 (85.33) |  |
| Female, n (%) | 129 (94.16) | 466 (58.47) | <0.001 |
| Age, median (IQR) | 41.07 (32.68- | 46.75 (36.84- | 0.001 |
|  | 49.95) | 59.27) |  |
| BAME, n (%) | 2 (1.46) | 10 (1.25) | 0.844 |
| In a relationship, n (%) | 34 (24.82) | 150 (18.82) | 0.103 |
| Death during follow-up | 9 (6.57) | 80 (10.04) | 0.202 |
| **Lithium exposure characteristics** |  |  |  |
| Lithium treatment duration (years), median (IQR) | 1.23 (0.37-2.66) | 1.23 (0.44-3.10) | 0.926 |
| Ever lithium toxic (>1.5mmol/L), n (%) | * | 37 (1.68) | 0.048 |
| Follow-up after stopping lithium (years), median (IQR) | 3.43 (0.42-7.36) | 4.44 (0.81-8.31) | 0.165 |
| **Renal function characteristics** |  |  |  |
| eGFR tests during follow-up, median (IQR) | 11 (6-21) | 14 (7-24) | 0.031 |
| Baseline eGFR (mL/min/1.73m<sup>2</sup>), median (IQR) | 66 (63-71) | 83 (74-94) | <0.001 |
| Developed CKD stage 3a or more severe (eGFR<60 mL/min/1.73m<sup>2</sup>) | 117 (85.40) | 112 (14.05) | <0.001 |
| Developed CKD stage 3b or more severe (eGFR<45 mL/min/1.73m<sup>2</sup>) | 14 (10.22) | * | <0.001 |
| **Pre-lithium mental health characteristics** |  |  |  |
| Depression, n (%) | 118 (86.13) | 634 (79.55) | 0.072 |
| Anxiety, n (%) | 396 (49.69) | 74 (54.01) | 0.349 |
| Psychosis, n (%) | 37 (27.01) | 179 (22.46) | 0.244 |
| Stress, n (%) | 25 (18.25) | 138 (17.31) | 0.790 |
| Self-harm, n (%) | 40 (29.20) | 185 (23.21) | 0.130 |
| Disturbed sleep, n (%) | 49 (35.77) | 202 (25.35) | 0.011 |
| Illness duration (years), median (IQR) | 7.73 (2.55-13.80) | 7.65 (2.71-15.48) | 0.926 |
| **Pre-lithium physical health characteristics** |  |  |  |
| Hypertension, n (%) | 15 (10.95) | 165 (20.70) | 0.008 |
| Migraine, n (%) | 24 (17.52) | 79 (9.91) | 0.009 |
| Type II diabetes mellitus, n (%) | 10 (7.30) | 64 (8.03) | 0.770 |
| Thyroid disease |  |  |  |
| Hypothyroidism, n (%) | 15 (10.95) | 57 (7.15) | 0.124 |
| Hyperthyroidism, n (%) | * | 5 (0.63) | 0.353 |
| Calcium abnormalities |  |  |  |
| hypocalcaemia, n (%) | * | 9 (1.13) | 0.740 |
| hypercalcaemia, n (%) | * | * | 0.678 |
| Cholesterol abnormalities |  |  |  |
| High LDL, n (%) | 20 (14.60) | 164 (20.58) | 0.104 |
| Low HDL, n (%) | 10 (7.30) | 66 (8.28) | 0.698 |
| Asthma, n (%) | 30 (21.90) | 131 (16.44) | 0.118 |
| Chronic obstructive pulmonary disease, n (%) | 36 (26.28) | 162 (20.33) | 0.115 |
| Anaemia, n (%) | 12 (8.76) | 30 (3.76) | 0.009 |
| Peptic ulcer, n (%) | 6 (4.38) | 27 (3.39) | 0.561 |
| Coronary heart disease, n (%) | * | 46 (5.77) | 0.035 |
| Liver disease, n (%) | * | 10 (1.25) | 0.138 |
| Neurological disorders, n (%) | 10 (7.30) | 45 (5.65) | 0.448 |
| Rheumatoid arthritis, n (%) | * | 17 (2.13) | 0.606 |
| Weight loss, n (%) | 5 (3.65) | 25 (3.14) | 0.753 |
| Ever eGFR<60 mL/min/1.73m2, n (%) | * | 105 (13.17) | <0.001 |
| **Health behaviours** |  |  |  |
| Smoking status, n (%) |  |  | 0.048 |
| Never smoked | 55 (40.15) | 297 (37.26) |  |
| Current smoker | 65 (47.45) | 329 (41.28) |  |
| Ex-smoker | 17 (12.41) | 171 (21.46) |  |
| Body mass index, n (%) |  |  | 0.259 |
| Underweight | * | 9 (1.13) |  |
| Healthy weight | 58 (42.34) | 321 (40.28) |  |
| Overweight | 34 (24.82) | 257 (32.25) |  |
| Obese | 42 (30.66) | 210 (26.35) |  |
| Cannabis use, n (%) | * | 13 (1.63) | 0.297 |
| Other substance misuse, n (%) | * | 21 (2.63) | 0.849 |
| Alcohol misuse, n (%) | 29 (21.17) | 111 (13.93) | 0.028 |
| **Other drug treatment** |  |  |  |
| Antipsychotic previously, n (%) | 73 (53.28) | 432 (54.20) | 0.842 |
| Mood stabiliser previously, n (%) | 38 (27.74) | 156 (19.57) | 0.030 |
| SSRI previously, n (%) | 43 (31.39) | 211 (26.47) | 0.233 |
| TCA previously, n (%) | 35 (25.55) | 193 (24.22) | 0.737 |
| Other antidepressant previously, n (%) | 79 (57.66) | 391 (49.06) | 0.063 |

\*n<5 individuals

**Figure 3.**
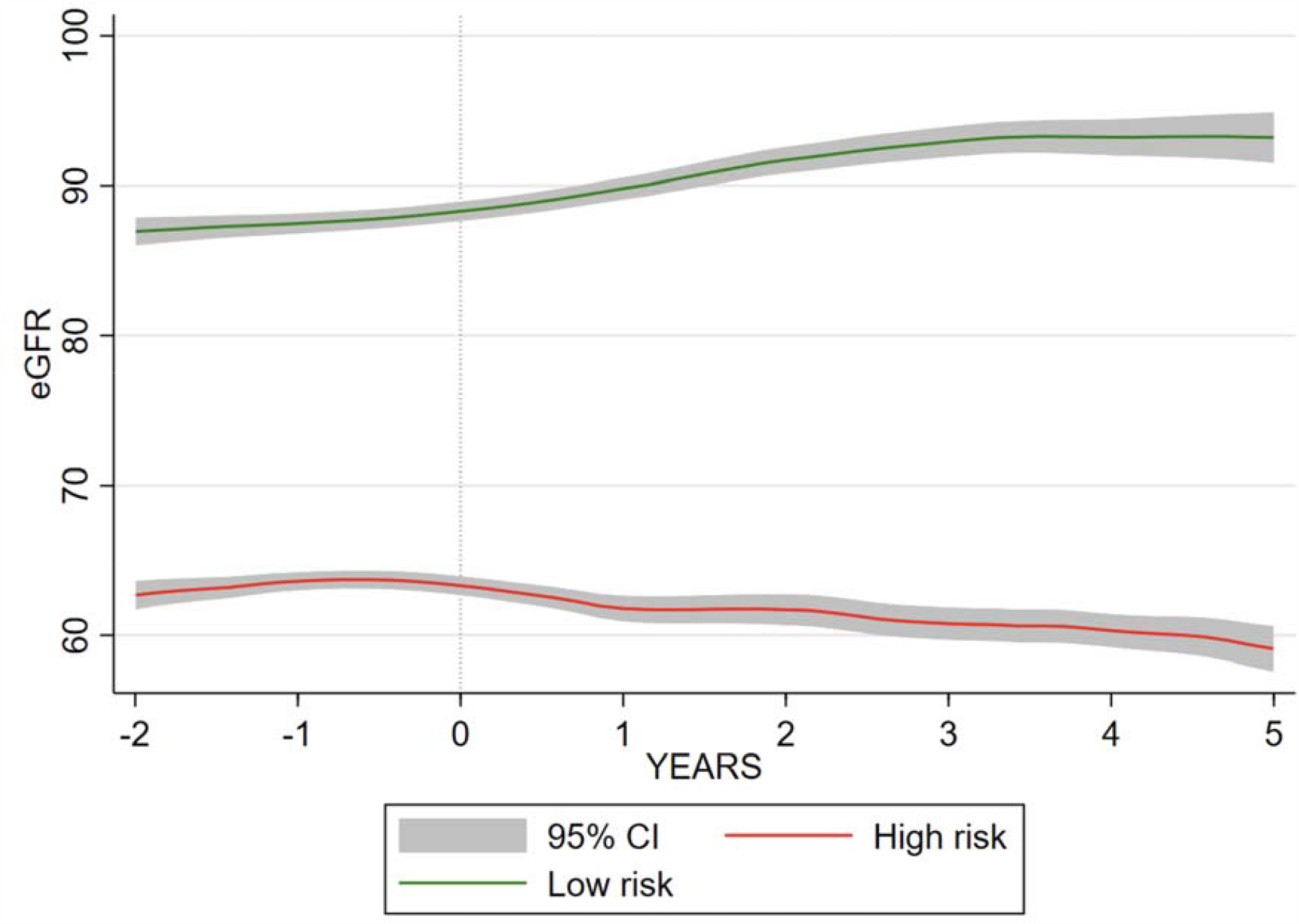
High-risk and low-risk eGFR trajectory in relation to stopping lithium in Gold

As with the Aurum cohort, patients in the high-risk group were more likely to be female, be younger, have a lower eGFR before starting lithium and less likely to have a prior record of eGFR<60 mL/min/1.73m^2^. High-risk individuals were also more likely to experience migraine. Unlike the Aurum cohort, the high-risk group were more likely to have anaemia and less likely to have hypertension. There was no between group difference for lithium duration and lithium toxicity was potentially more common in the low-risk group.

We predicted high-risk group membership using the model generated in the Aurum Data set. The ROC area was 0.879 (95%CI 0.853-0.904) (Table 2, Figure 4). At the empirical optimal cut-point defined in the development dataset, the model had a sensitivity of 0. 91 (0.84-0.97) and a specificity of 0.74 (95% CI 0.67-0.82) The Brier score was 0.0967. This gave a predictive accuracy of 76.55%. However, the simpler models also predicted high-risk group membership similarly well: 3-variable KFRE ROC area 0.870 (95%CI 0.841-0.898), 3-variable elastic net ROC area 0.888 (95%CI 0.864-0.912)(Box 1). The calibration plot suggested that the model performs well up to a probability of 0.60 at the 95% confidence level (Figure 5).

**Figure 4.**
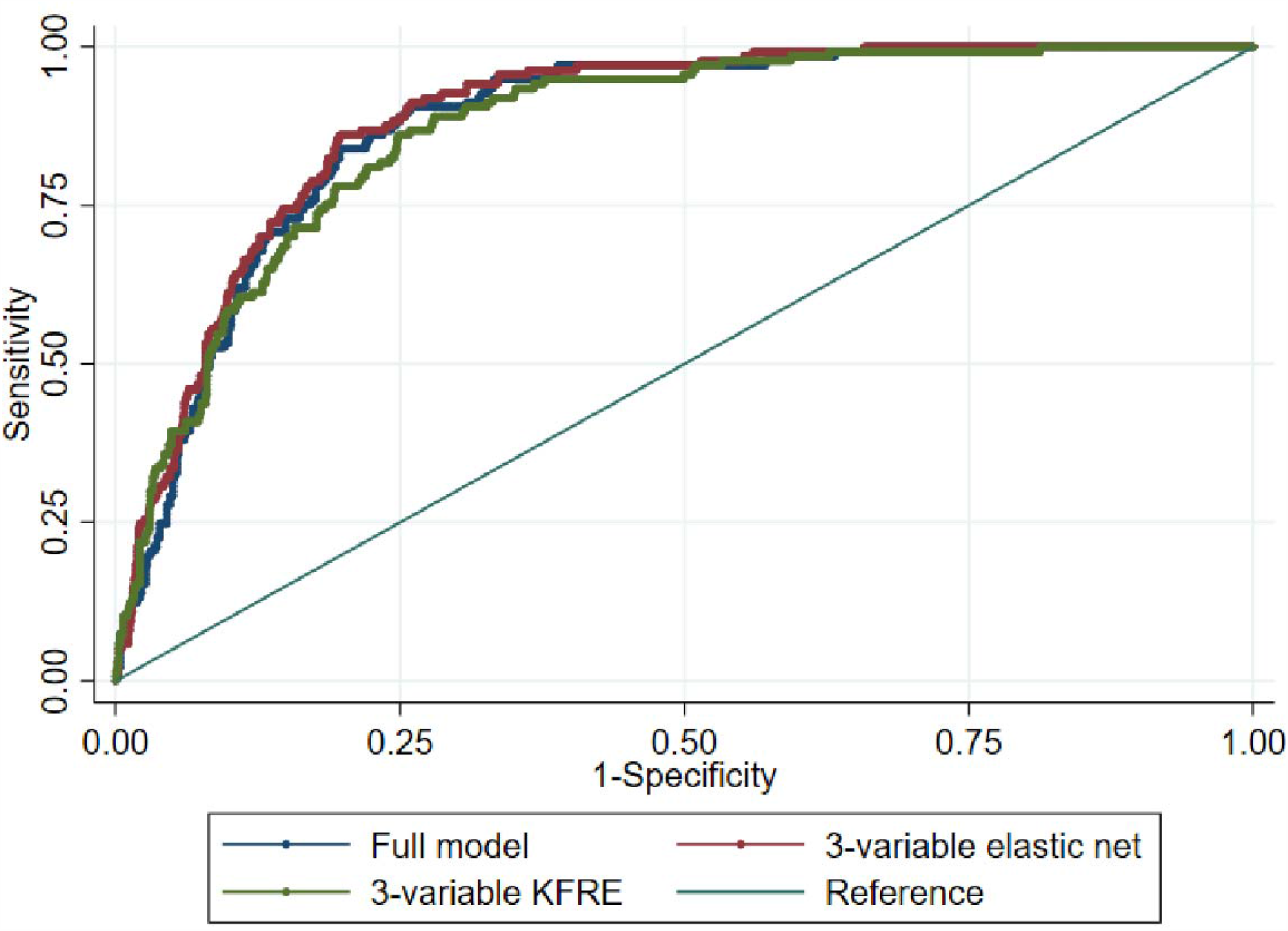
Sensitivity vs specificity of the high-risk trajectory prediction model in Gold

**Figure 5.**
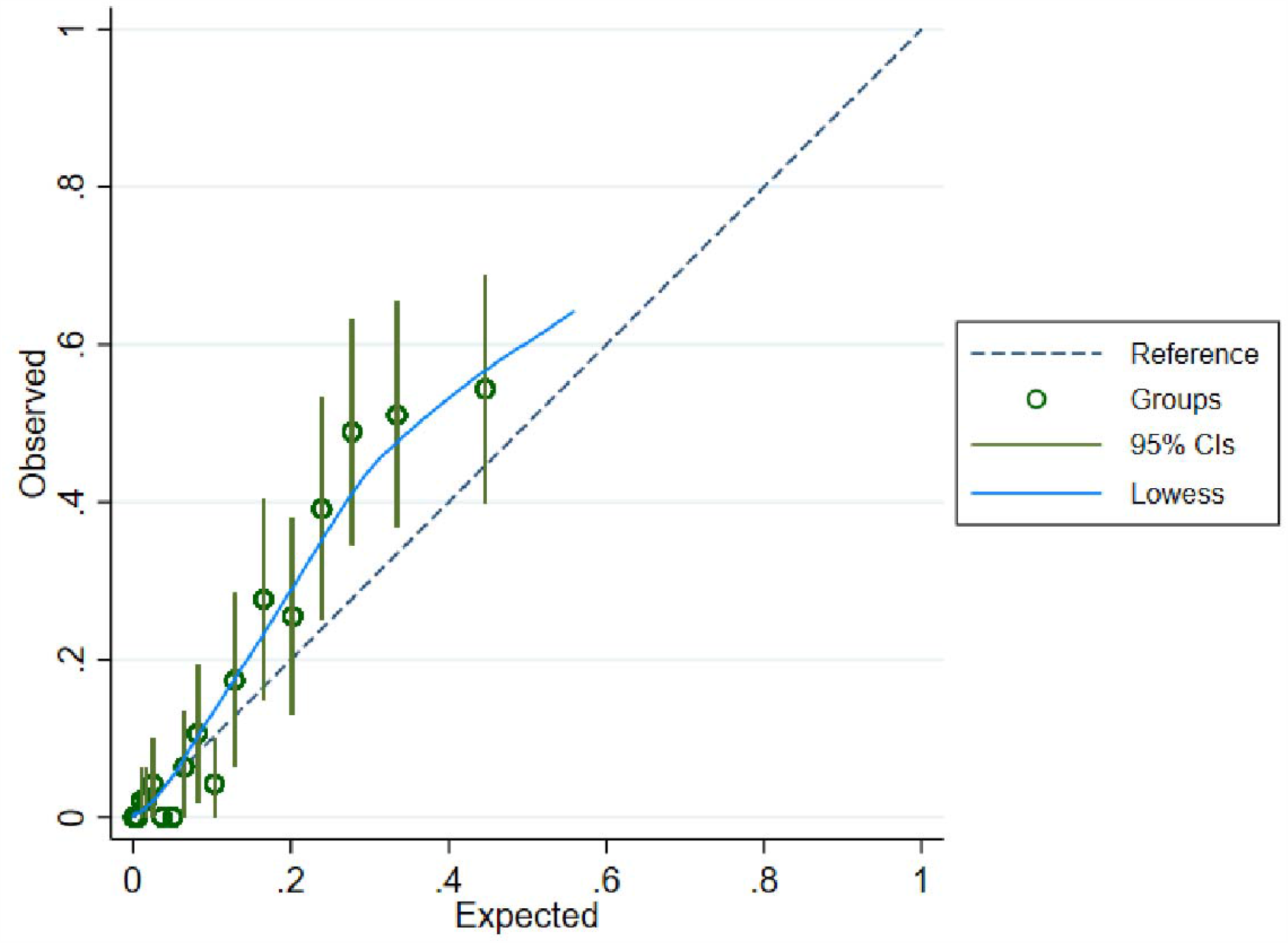
Calibration plot for Gold data set (20 groups across risk spectrum)

#### Box 1. 3-variable equation for prediction of high-risk group membership

Risk score = 4.621 - (0.717 x eGFR) - (0.616 x sex^†^) - (0.008 x age)

probability of high-risk group membership = exp (risk score)/ (1+exp (risk score))

^†^where sex=1 if male and sex=0 if female

We also predicted CKD 3b or more severe using these models: ROC area 0.849 (95%CI 0.792-0.905), ROC area 0.865 (95%CI 0.808-0.922), ROC area 0.858 (95%CI 0.792-0.922) using the full model, 3-variable elastic net and 3-variable KFRE respectively (Table 4).

**Table 4.** Prediction of CKD stage 3b or more severe

| MODEL | ROC area (95%CI) | Sensitivity (95%CI) | Specificity (95%CI) | Accuracy, % |
| --- | --- | --- | --- | --- |
| **Development (Aurum)** |  |  |  |  |
| Full | 0.700 (0.617-0.783) | 0.71 (0.52-0.90) | 0.66 (0.47-0.85) | 66.12 |
| 3-variable KFRE | 0.627 (0.543-0.710) | 0.68 (0.51-0.86) | 0.61 (0.48-0.74) | 61.18 |
| 3-variable elastic net | 0.678 (0.598-0.758) | 0.58 (0.37-0.78) | 0.73 (0.48-0.98) | 72.64 |
| **Validation (Gold)** |  |  |  |  |
| Full | 0.849 (0.792-0.905) | 1.00 (0.96-1.00) | 0.67 (0.57-0.76) | 67.49 |
| 3-variable KFRE | 0.858 (0.792-0.922) | 0.86 (0.70-1.00) | 0.78 (0.63-0.93) | 78.12 |
| 3-variable elastic net | 0.865 (0.808- | 1.00 (0.92- | 0.66 (0.57-0.75) | 66.51 |
|  | 0.921) | 1.00) |  |  |

The decision curve analysis showed that all 3 of these models were superior to classifying everyone as high-risk or low-risk between a threshold probability of 0.10 and 0.80 and there was little difference between them (Figure 6).

**Figure 6.**
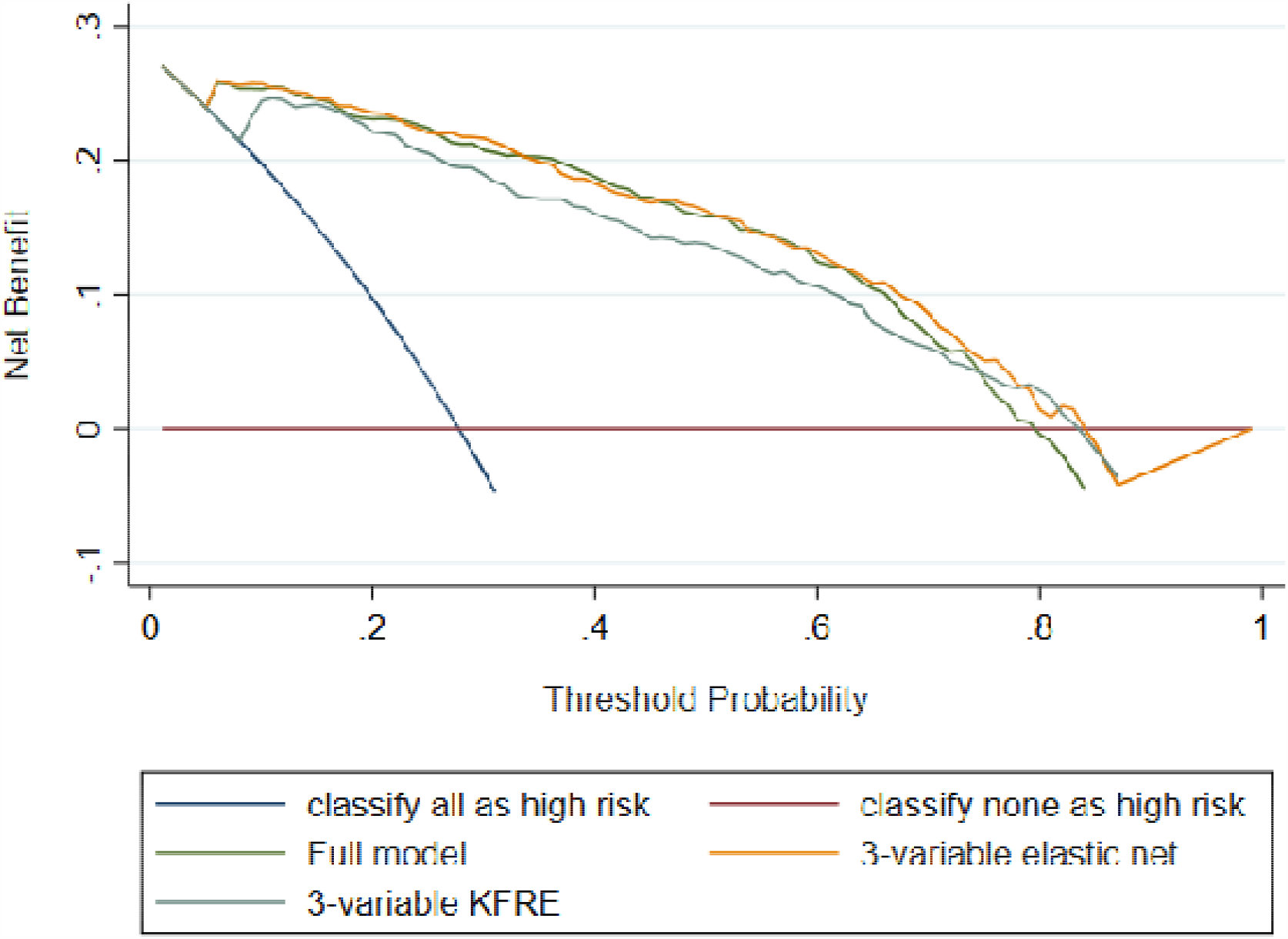
Decision curve analysis for Gold data set

### Post hoc supplementary analysis

In 668 patients with a baseline eGFR ≥90 mL/min/1.73m^2^ a two-group cubic trajectory model minimised the BIC (642.39) with 120 patients (17.96%) in the high-risk group (Table 5, Figure 7). CKD stage 3a or more severe and stage 3b or more severe were more common in the high-risk group. Individuals in the high-risk group were again more likely to be female, be younger and have a lower eGFR before starting lithium. They were more likely to be current smokers. We did not observe any of the other between group differences present in the Aurum or Gold trajectory groups.

**Table 5.** Characteristics of lithium prescribed patients by risk group in patients with baseline eGFR ≥90mL/min/1.73m^2^

| Feature | High-risk | Low-risk | P-value |
| --- | --- | --- | --- |
| **Patient characteristics** |  |  |  |
| Total, N (%) | 120 (17.96) | 548 (82.04) |  |
| Female, n (%) | 65 (54.17) | 158 (28.83) | <0.001 |
| Age, median (IQR) | 45.48 (34.99-55.58) | 52.18 (42.03-65.93) | <0.001 |
| BAME, n (%) | * | 15 (2.74) | 0.500 |
| In a relationship, n (%) | 23 (19.17) | 82 (14.96) | 0.252 |
| Death during follow-up | 13 (10.83) | 94 (17.15) | 0.087 |
| **Lithium exposure characteristics** |  |  |  |
| Lithium treatment duration (years), median (IQR) | 1.23 (0.54-3.07) | 1.61 (0.61-3.84) | 0.087 |
| Ever lithium toxic (>1.5mmol/L), n (%) | 7 (5.83) | 36 (6.57) | 0.867 |
| Follow-up after stopping lithium (years), median (IQR) | 8.01 (2.68-12.08) | 4.77 (0.97-9.77) | 0.001 |
| **Renal function characteristics** |  |  |  |
| Baseline eGFR (mL/min/1.73m<sup>2</sup>), median (IQR) | 95 (92-100) | 100 (94-108) | <0.001 |
| Developed CKD stage 3a or more severe (eGFR<60 mL/min/1.73m<sup>2</sup>) | 34 (28.33) | * | <0.001 |
| Developed CKD stage 3b or more severe (eGFR<45 mL/min/1.73m<sup>2</sup>) | * | * | <0.001 |
| **Pre-lithium mental health characteristics** |  |  |  |
| Depression, n (%) | 97 (80.83) | 423 (77.19) | 0.384 |
| Anxiety, n (%) | 34 (28.33) | 188 (34.31) | 0.208 |
| Psychosis, n (%) | 26 (21.67) | 133 (24.27) | 0.544 |
| Stress, n (%) | 14 (11.67) | 71 (12.96) | 0.701 |
| Self-harm, n (%) | 24 (20.00) | 84 (15.33) | 0.208 |
| Disturbed sleep, n (%) | 24 (20.00) | 113 (20.62) | 0.879 |
| Illness duration (years), median (IQR) | 7.43 (1.81-12.87) | 7.26 (2.53-17.79) | 0.687 |
| **Pre-lithium physical health characteristics** |  |  |  |
| Hypertension, n (%) | 20 (16.67) | 140 (25.55) | 0.039 |
| Migraine, n (%) | 9 (7.50) | 35 (6.39) | 0.656 |
| Type II diabetes mellitus, n (%) | 7 (5.83) | 58 (10.58) | 0.112 |
| Thyroid disease |  |  |  |
| Hypothyroidism, n (%) | * | 24 (4.38) | 0.604 |
| Hyperthyroidism, n (%) | * | * | 0.507 |
| Calcium abnormalities |  |  |  |
| hypocalcaemia, n (%) | * | * | 0.934 |
| hypercalcaemia, n (%) | * | 6 (1.09) | 0.799 |
| Cholesterol abnormalities |  |  |  |
| High LDL, n (%) | 29 (24.17) | 97 (17.70) | 0.101 |
| Low HDL, n (%) | 12 (10.00) | 46 (8.39) | 0.572 |
| Asthma, n (%) | 25 (20.83) | 75 (13.69) | 0.047 |
| Chronic obstructive pulmonary disease, n (%) | 29 (24.17) | 100 (18.25) | 0.137 |
| Anaemia, n (%) | * | 20 (3.65) | 0.532 |
| Peptic ulcer, n (%) | * | 16 (2.92) | 0.189 |
| Coronary heart disease, n (%) | 5 (4.17) | 49 (8.94) | 0.082 |
| Liver disease, n (%) | * | 8 (1.46) | 0.590 |
| Neurological disorders, n (%) | 6 (5.00) | 41 (7.48) | 0.4336 |
| Rheumatoid arthritis, n (%) | 5 (4.17) | 11 (2.01) | 0.161 |
| Weight loss, n (%) | * | 11 (2.01) | 0.381 |
| Ever eGFR<60 mL/min/1.73m2, n (%) | 16 (13.33) | 105 (19.16) | 0.133 |
| **Health behaviours** |  |  |  |
| Smoking status, n (%) |  |  | 0.001 |
| Never smoked | 26 (21.67) | 197 (35.95) |  |
| Current smoker | 62 (51.67) | 193 (35.22) |  |
| Ex-smoker | 32 (26.67) | 158 (28.83) |  |
| Body mass index, n (%) |  |  | 0.534 |
| Underweight | * | 9 (1.28) |  |
| Healthy weight | 47 (39.17) | 227 (41.42) |  |
| Overweight | 36 (30.00) | 187 (34.12) |  |
| Obese | 35 (29.17) | 127 (23.18) |  |
| Cannabis use, n (%) | * | 6 (1.09) | 0.799 |
| Other substance misuse, n (%) | 7 (5.83) | 19 (3.47) | 0.225 |
| Alcohol misuse, n (%) | 8 (6.67) | 47 (8.58) | 0.491 |
| **Other drug treatment** |  |  |  |
| Antipsychotic previously, n (%) | 65 (54.17) | 269 (49.09) | 0.314 |
| Mood stabiliser previously, n (%) | 34 (28.33) | 117 (21.35) | 0.098 |
| SSRI previously, n (%) | 31 (25.83) | 126 (22.99) | 0.506 |
| TCA previously, n (%) | 19 (15.83) | 120 (21.90) | 0.138 |
| Other antidepressant previously, n (%) | 16 (13.33) | 60 (10.95) | 0.456 |

\*n<5 individuals

**Figure 7.**
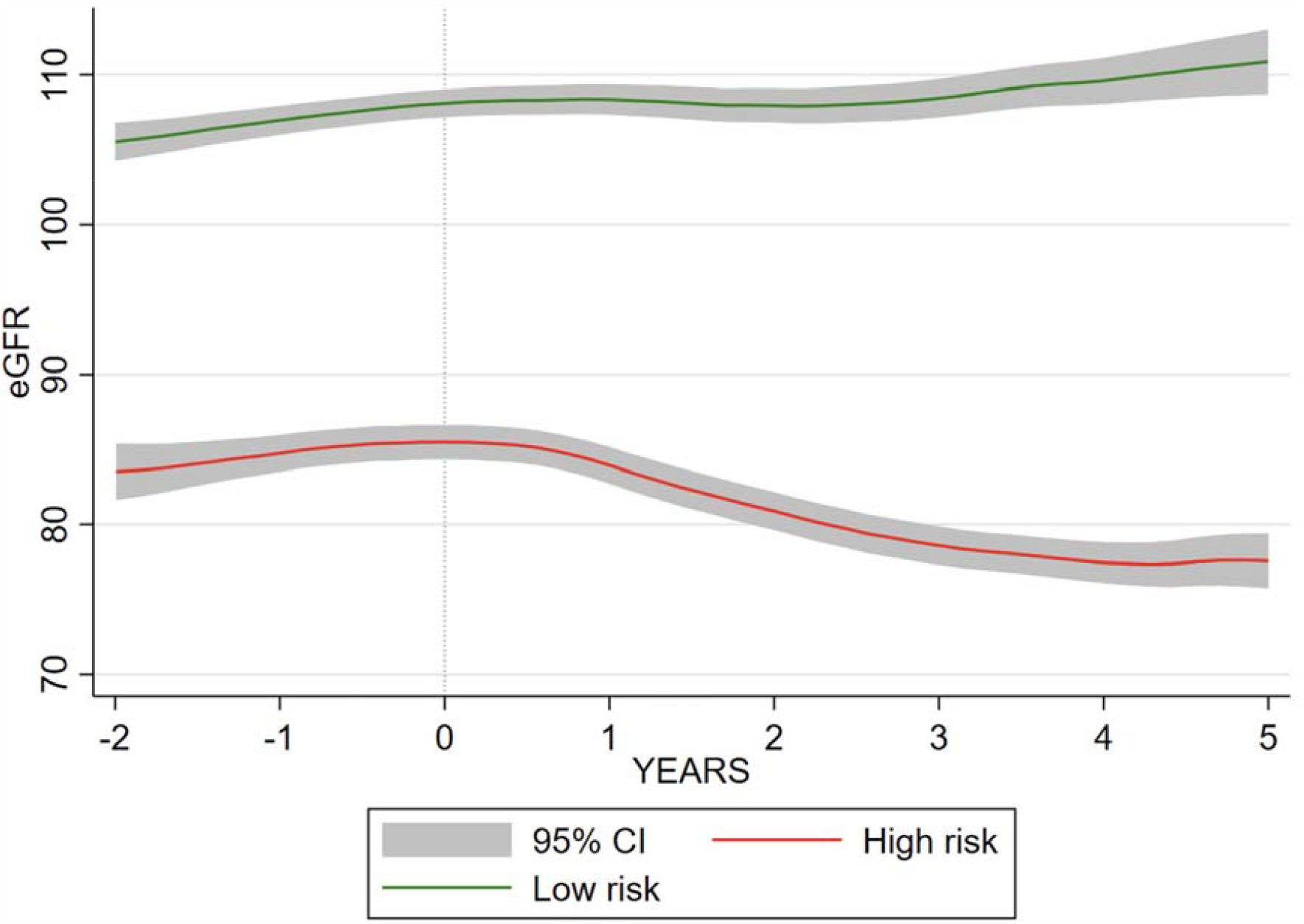
High-risk and low-risk eGFR trajectory in relation to stopping lithium in patients with baseline eGFR ≥90mL/min/1.73m^2^

In this reduced dataset our full model was better at predicting high-risk group membership than the 3-variable KFRE model (ROC area 0.725; 95%CI 0.675-0.776 vs ROC area 0.667; 95%CI 0.617-0.716; p-value for equality=0.0018), but not the 3-varible elastic net model (ROC area 0.729; 95%CI 0.679-0.779; p-value for equality 0.5846) (Table 6, Figure 8).

**Table 6.**
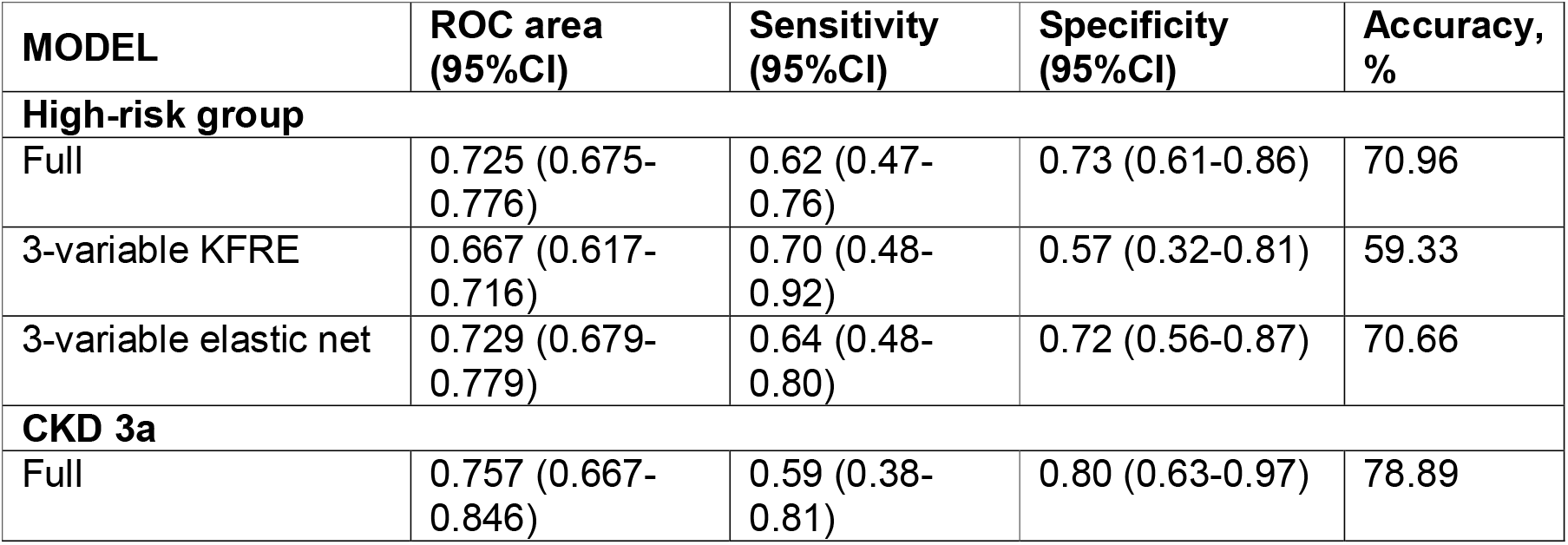
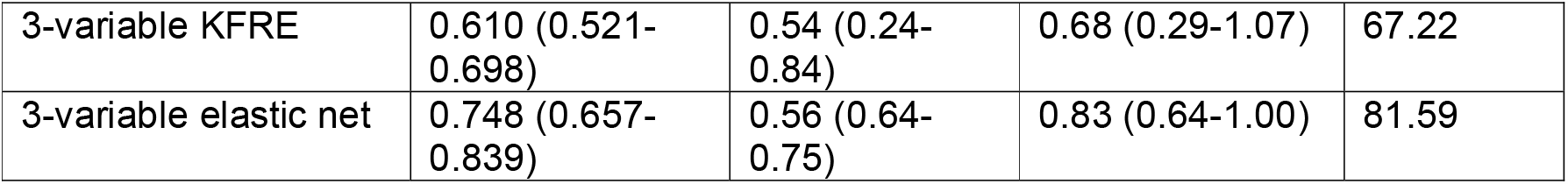
Prediction in individuals with baseline eGFR≥90mL/min/1.73m^2^

| MODEL | ROC area (95%CI) | Sensitivity (95%CI) | Specificity (95%CI) | Accuracy, % |
| --- | --- | --- | --- | --- |
| **High-risk group** |  |  |  |  |
| Full | 0.725 (0.675-0.776) | 0.62 (0.47-0.76) | 0.73 (0.61-0.86) | 70.96 |
| 3-variable KFRE | 0.667 (0.617-0.716) | 0.70 (0.48-0.92) | 0.57 (0.32-0.81) | 59.33 |
| 3-variable elastic net | 0.729 (0.679-0.779) | 0.64 (0.48-0.80) | 0.72 (0.56-0.87) | 70.66 |
| **CKD 3a** |  |  |  |  |
| Full | 0.757 (0.667-0.846) | 0.59 (0.38-0.81) | 0.80 (0.63-0.97) | 78.89 |
| 3-variable KFRE | 0.610 (0.521-0.698) | 0.54 (0.24-0.84) | 0.68 (0.29-1.07) | 67.22 |
| 3-variable elastic net | 0.748 (0.657-0.839) | 0.56 (0.64-0.75) | 0.83 (0.64-1.00) | 81.59 |

**Figure 8.**
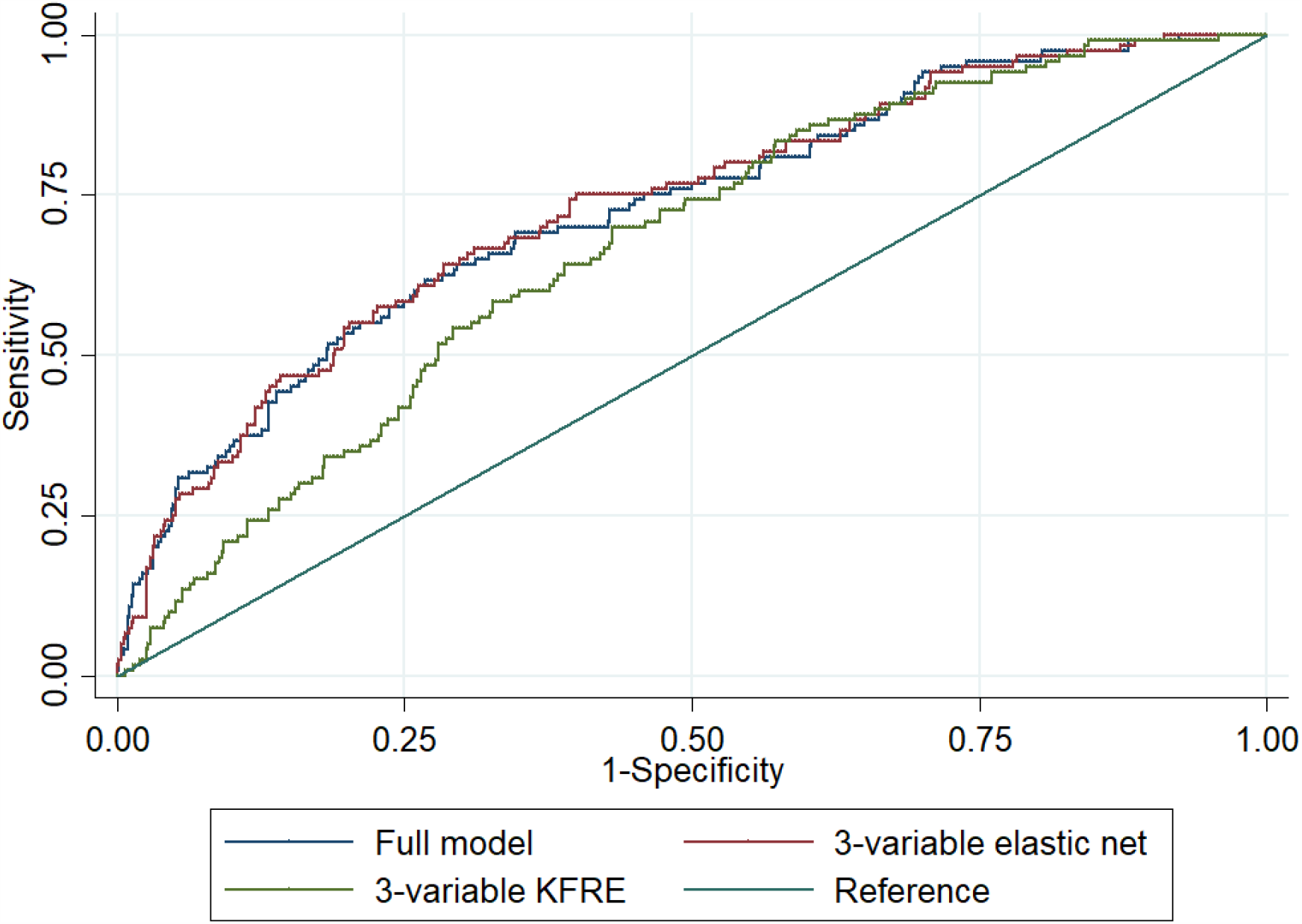
Comparison of ROC areas between full model, 3-variable model and 3-variable KFRE model

## Discussion

As far as we are aware, this is the first model developed to predict high-risk of future impaired renal function in people with bipolar disorder prescribed lithium. We used a large representative sample of people with bipolar disorder initiated on lithium and followed up for up for a median of 7.10 years (IQR 3.85-11.36). It is also the first study to use the two CPRD datasets, covering a large, representative sample of the UK population to develop a prediction model and provide external validation (external validation is a vital step often lacking in such studies).

Because of the rarity of renal failure, and the varied follow-up time and eGFR recording frequency in EHRs, we sought to identify approximately 10% of individuals prescribed lithium who were at highest risk of deteriorating renal function, defined by the trajectory of their serial eGFR measurements. Using group-based trajectory analysis we identified high-risk groups independently in the Aurum and Gold cohorts. In both cases approximately 85% of those categorised as high-risk developed CKD stage 3a or more severe compared to approximately 15% in the low-risk groups. Approximately 10% of those identified as high-risk developed CKD stage 3b or more severe, compared to <1% in the low-risk group. A number of features differed between the high-risk and low-risk groups in both cohorts. Those in the high-risk groups were more likely to be female, younger, more likely to have a lower eGFR before starting lithium, more likely to experience migraine and less likely to have a prior record of eGFR<60 mL/min/1.73m^2^. These CKD risk factors have been previously identified. CKD is more common in women (24), and this has been shown in lithium users specifically (25). Younger women appear to be at particular risk (25). Low baseline eGFR increases risk of CKD in the general population (26). Migraine is not commonly thought of as a risk factor for CKD, but has been identified as such in one study, especially in younger age groups (27). Migraine is highly comorbid with bipolar disorder (28) and it may also be a proxy for medication use which could impair renal function. In both cohorts, there was no association between duration of lithium treatment or lithium toxicity (which was rare) and high-risk group membership.

Our model, developed in CPRD Aurum to predict whether individuals were in a high-risk group for deteriorating renal function during treatment with lithium for bipolar disorder, had excellent discrimination in the CPRD Gold cohort (ROC area 0.879). However, simple models only including sex, age and baseline eGFR performed similarly well (3-variable KFRE ROC area 0.870, 3-variable elastic net ROC area 0.888), all with similar levels of accuracy (>75%). In the external validation data set, our model designed to predict high-risk trajectory predicted CKD stage 3b or more severe (eGFR <45 mL/min/1.73m^2^) with a ROC area 0.849. Again, simple models performed well, with the 3-variable KFRE having the highest accuracy (78%).

When we reduced our cohort to those starting lithium with an eGFR ≥90 mL/min/1.73m^2^ Our full model and 3-variable elastic net model performed better than the KFRE. However, our sample was too small to complete external validation of these new models.

Calculating risk of renal impairment could help clinicians and patients better understand the potential for adverse effects of lithium and guide shared decision making. Given our findings, simple risk calculators should be used in clinical practice at the decision to commence lithium and when eGFR is regularly measured, and should be added to lithium monitoring protocols. Risk could be calculated via the KFRE or our 3-varaible elastic net model (Box 1), which performs better than the 3-variable KFRE when eGFR ≥90 mL/min/1.73m^2^. We did not find predictors of poor renal function that were specific to lithium treated patients.

### Strengths and Limitations

Our large, population-based longitudinal sample avoided selection bias and is generalisable and representative. Our group-based trajectory approach avoided issues with differential follow-up time and potential surveillance bias. Our use of elastic net allowed us to build a parsimonious prediction model from a large number of potential features.

The study has a number of limitations. Instead of using precise eGFR values to define outcome we split individuals into those with a high-risk and low-risk trajectory. We forced the group-based trajectory model to identify >10% of individuals prescribed lithium who were at highest risk of developing impaired renal function. A more useful model clinically would be to predict true renal failure requiring intervention; however, this was too rare in our cohort (2% developed CKD stage 3b or more severe), suggesting it is uncommon in modern clinical practice. We included a large number of potential predictors. However, we may not have included all important features in our elastic net model. Some variables of interest, such as proteinuria, were poorly recorded and biased by diabetes diagnosis.

It is possible that we failed to identify people previously exposed to lithium, but we attempted to limit this by ensuring patients had at least a year of follow-up at the same primary care practice before their first identified lithium prescription. In most cases this would also include the uploading of historical records to the EHR. It may be that we excluded people with longer lithium exposure because of this inclusion criteria, and these excluded patients may be at higher risk of CKD, however our cohort did include indivuduals with up to 17 years of lithium exposure. Patients could also be misclassified in terms of different features included in the model. However, our intention was to build a model based on what is already known about the patient from the EHR. Misclassification may be more likely for some features (for example, in a relationship) than others (for example, chronic obstructive pulmonary disease).

We initially planned to develop a model for individuals with essentially normal renal function (eGFR ≥60 mL/min/1.73m^2^). Although the discrimination and calibration of the model was good, a simple model based on baseline eGFR, age and sex performed just as well.

## Conclusion

We developed a model for predicting, at lithium initiation, individuals at high-risk of a poor trajectory of renal function using serial eGFR measurements. We externally validated this model, which had excellent discrimination and good calibration. CKD stage 3b or more severe occurred in 2% of the population across the two cohorts. Whilst this is worrying, it means that the vast majority of patients treated with lithium do not develop renal failure, and those at risk can be identified prior to initiating lithium using their age, sex and baseline eGFR.

## Data Availability

Data is avaiable via the Clinical Practice Research Datalink (https://www.cprd.com/) following appropriate ethical approval.

## References

1. Barroilhet SA, Ghaemi SN. When and how to use Lithium. Acta Psychiatrica Scandinavica. 2020.

2. Tondo L, Alda M, Bauer M, Bergink V, Grof P, Hajek T, et al. Clinical use of lithium salts: guide for users and prescribers. International journal of bipolar disorders. 2019;7(1):16.

3. Gitlin M. Lithium side effects and toxicity: prevalence and management strategies. International journal of bipolar disorders. 2016;4(1):1–10.

4. McKnight RF, Adida M, Budge K, Stockton S, Goodwin GM, Geddes JR. Lithium toxicity profile: a systematic review and meta-analysis. The Lancet. 2012;379(9817):721–8.

5. Hayes JF, Marston L, Walters K, Geddes JR, King M, Osborn DP. Adverse renal, endocrine, hepatic, and metabolic events during maintenance mood stabilizer treatment for bipolar disorder: a population-based cohort study. PLoS medicine. 2016;13(8).

6. Iwagami M, Mansfield KE, Hayes JF, Walters K, Osborn DP, Smeeth L, et al. Severe mental illness and chronic kidney disease: a cross-sectional study in the United Kingdom. Clinical epidemiology. 2018;10:421.

7. Kessing LV, Gerds TA, Feldt-Rasmussen B, Andersen PK, Licht RW. Use of lithium and anticonvulsants and the rate of chronic kidney disease: a nationwide population-based study. JAMA psychiatry. 2015;72(12):1182–91.

8. Gupta S, Khastgir U. Drug information update. Lithium and chronic kidney disease: debates and dilemmas. BJPsych Bulletin. 2017;41(4):216–20.

9. Kazancioğlu R. Risk factors for chronic kidney disease: an update. Kidney international supplements. 2013;3(4):368–71.

10. Lim CC, Chee ML, Cheng C-Y, Kwek JL, Foo M, Wong TY, et al. Simplified end stage renal failure risk prediction model for the low-risk general population with chronic kidney disease. PloS one. 2019;14(2):e0212590.

11. Ramspek CL, de Jong Y, Dekker FW, van Diepen M. Towards the best kidney failure prediction tool: a systematic review and selection aid. Nephrology Dialysis Transplantation. 2019.

12. Johnson ES, Thorp ML, Platt RW, Smith DH. Predicting the risk of dialysis and transplant among patients with CKD: a retrospective cohort study. American journal of kidney diseases. 2008;52(4):653–60.

13. Rigatto C, Sood MM, Tangri N. Risk prediction in chronic kidney disease: pitfalls and caveats. Current opinion in nephrology and hypertension. 2012;21(6):612–8.

14. Tangri N, Grams ME, Levey AS, Coresh J, Appel LJ, Astor BC, et al. Multinational assessment of accuracy of equations for predicting risk of kidney failure: a meta-analysis. Jama. 2016;315(2):164–74.

15. Herrett E, Gallagher AM, Bhaskaran K, Forbes H, Mathur R, Van Staa T, et al. Data resource profile: clinical practice research datalink (CPRD). International journal of epidemiology. 2015;44(3):827–36.

16. Wolf A, Dedman D, Campbell J, Booth H, Lunn D, Chapman J, et al. Data resource profile: Clinical practice research datalink (cprd) aurum. International journal of epidemiology. 2019;48(6):1740–g.

17. Matsushita K, Mahmoodi BK, Woodward M, Emberson JR, Jafar TH, Jee SH, et al. Comparison of risk prediction using the CKD-EPI equation and the MDRD study equation for estimated glomerular filtration rate. Jama. 2012;307(18):1941–51.

18. Nagin DS, Odgers CL. Group-based trajectory modeling in clinical research. Annual review of clinical psychology. 2010;6:109–38.

19. Nylund KL, Asparouhov T, Muthén BO. Deciding on the number of classes in latent class analysis and growth mixture modeling: A Monte Carlo simulation study. Structural equation modeling: A multidisciplinary Journal. 2007;14(4):535–69.

20. Zou H, Hastie T. Regularization and variable selection via the elastic net. Journal of the royal statistical society: series B (statistical methodology). 2005;67(2):301–20.

21. Tangri N, Stevens LA, Griffith J, Tighiouart H, Djurdjev O, Naimark D, et al. A predictive model for progression of chronic kidney disease to kidney failure. Jama. 2011;305(15):1553–9.

22. Nattino G, Finazzi S, Bertolini G. A new calibration test and a reappraisal of the calibration belt for the assessment of prediction models based on dichotomous outcomes. Statistics in medicine. 2014;33(14):2390–407.

23. StataCorp L. Stata statistical software: release 16. College Station, TX. 2019.

24. Carrero JJ, Hecking M, Chesnaye NC, Jager KJ. Sex and gender disparities in the epidemiology and outcomes of chronic kidney disease. Nature reviews Nephrology. 2018;14(3):151.

25. Shine B, McKnight RF, Leaver L, Geddes JR. Long-term effects of lithium on renal, thyroid, and parathyroid function: a retrospective analysis of laboratory data. The Lancet. 2015;386(9992):461–8.

26. Major RW, Shepherd D, Medcalf JF, Xu G, Gray LJ, Brunskill NJ. The Kidney Failure Risk Equation for prediction of end stage renal disease in UK primary care: An external validation and clinical impact projection cohort study. PLoS medicine. 2019;16(11):e1002955.

27. Weng S-C, Wu C-L, Kor C-T, Chiu P-F, Wu M-J, Chang C-C, et al. Migraine and subsequent chronic kidney disease risk: a nationwide population-based cohort study. BMJ open. 2017;7(12):e018483.

28. Leo RJ, Singh J. Migraine headache and bipolar disorder comorbidity: A systematic review of the literature and clinical implications. Scandinavian Journal of Pain. 2016;11:136–45.

